# Prevalence of Visceral Leishmaniasis among Wildlife Rangers in Dinder National Park, Sudan

**DOI:** 10.1101/2024.06.24.24309386

**Authors:** Bashir Ibrahim, Mohammed Elmadani, Hemeda Sahar, Mogahid Gadallh, A. Abdallh, Abd Elbasit Elawad Ahmed

**Author notes:** Corresponding Authors: Mohammed Elmadani: Lecturer of Public Health and Epidemiology, University of El Imam El Mahdi, Faculty of Public Health, Department of Epidemiology, Kosti, Sudan, 27711.

## Abstract

**Background:** Visceral leishmaniasis (VL) is a significant public health concern in Sudan, particularly among populations exposed to vector-rich environments. This study aims to determine the prevalence of VL and associated risk factors among Wildlife Soldiers in Dinder National Park, Sudan.

**Methods:** A descriptive cross-sectional community-based study was conducted among 500 Wildlife Soldiers in Dinder National Park. Data were collected using a pre-prepared and pre-tested questionnaire covering demographic information, risk factors, and clinical signs and symptoms of VL. Data analysis involved descriptive statistics, Chi-square tests, and logistic regression to assess associations between VL prevalence and various risk factors.

**Results:** The prevalence of VL was found to be 27.6%. Significant associations were observed between VL prevalence and educational level, military rank, awareness of VL signs and symptoms, awareness of VL transmission methods, awareness of VL control measures, and the practice of sleeping under a mosquito net. Soldiers with higher educational levels, those who were aware of VL transmission and control measures, and those who slept under nets had significantly lower odds of contracting VL. Conversely, lower-ranking soldiers and those with less awareness had higher prevalence rates.

**Conclusion:** The high prevalence of VL among Wildlife Soldiers in Dinder National Park underscores the need for targeted public health interventions. Strategies should include enhancing educational programs, improving awareness of VL prevention and control measures, and ensuring better access to protective measures such as mosquito nets. Future research should focus on longitudinal studies, detailed environmental assessments, and intervention trials to further reduce the burden of VL in this high-risk population.

**Author Summary:** Visceral leishmaniasis (VL), or kala-azar, is a severe parasitic disease posing a significant health threat globally, including Sudan. This cross-sectional study investigates VL prevalence and associated risk factors among 500 Wildlife Soldiers in Dinder National Park, Sinnar State, Sudan. Data on demographics, risk factors, and VL signs were collected using a validated questionnaire. Findings reveal a 27.6% VL prevalence among soldiers, with significant associations noted for education level, military rank, VL awareness, transmission knowledge, and mosquito net use. Higher education and awareness correlated with lower VL rates, while lower-ranked and less-aware soldiers had higher prevalence. Targeted public health interventions are crucial to mitigate VL among Wildlife Soldiers, emphasizing education enhancement, awareness campaigns, and access to protective gear. Future research should focus on longitudinal studies and environmental assessments to refine VL control strategies in this high-risk population. This study enhances understanding of VL factors in Dinder National Park and advocates for robust public health initiatives to combat this disease effectively.

## Introduction

Visceral leishmaniasis (VL), also known as kala-azar, is a severe parasitic disease caused by protozoa of the *Leishmania* genus [1, 2]. It is transmitted through the bite of infected female phlebotomine sandflies and primarily affects internal organs such as the spleen, liver, and bone marrow[2, 3]. With an estimated 50,000 to 90,000 new cases annually, VL is a significant public health concern, particularly in tropical and subtropical regions where environmental conditions favor the sandfly vectors and the reservoirs of the parasite [2, 4–6] .

Sudan is one of the six countries contributing to more than 90% of global VL cases[1, 7]. The disease is endemic in the eastern and central regions, where it poses a substantial health burden[8–10]. Dinder National Park, located in southeastern Sudan, is a prominent conservation area that houses diverse wildlife and is a known habitat for sandflies [11, 12]. The interaction between wildlife, sandflies, and humans in this ecological setting creates a conducive environment for the transmission of VL [13–15].

Visceral leishmaniasis (VL) is a significant health concern in Sudan, particularly in the central and eastern regions [8, 10]. The prevalence of VL in these areas is high, with a particularly high incidence among children [16, 17]. The disease is associated with a range of clinical symptoms, including fever, pallor, weight loss, and splenomegaly [16]. Despite the availability of treatment, VL continues to pose a public health challenge in Sudan, with a high mortality rate [17].

Wildlife rangers working in Dinder National Park are at heightened risk of VL due to their prolonged exposure to the park’s environment. Rangers are essential for the protection and management of wildlife resources, yet their occupational activities often place them near the vector and reservoir hosts. Despite the critical role rangers play, there is a paucity of data on the prevalence of VL among this vulnerable population, limiting the implementation of targeted health interventions.

Understanding the prevalence of VL among wildlife rangers is vital for several reasons. First, it will highlight the occupational health risks associated with their work and the need for protective measures. Second, it will provide insights into the epidemiology of VL in a unique ecological niche, contributing to broader public health knowledge. Third, such data can inform policymakers and health authorities in developing effective strategies for disease prevention and control in similar high-risk groups.

This study aims to assess the prevalence of visceral leishmaniasis among wildlife rangers in Dinder National Park, Sudan. By investigating the extent of VL infection and associated risk factors in this population, we hope to provide evidence-based recommendations to improve the health and safety of wildlife rangers. Additionally, the findings may offer a model for addressing VL in other conservation areas and occupations with similar exposure risks.

Through this research, we seek to bridge the gap in the literature on occupational health risks of VL and emphasize the importance of integrated disease management approaches in endemic regions. This study underscores the need for continuous surveillance, enhanced diagnostic facilities, and comprehensive health education programs to mitigate the impact of VL on those most at risk.

## Materials and methods

This descriptive cross-sectional community-based study was conducted in Dinder National Park, Sinnar State, Sudan, using quantitative research methods to investigate the prevalence of Visceral Leishmaniasis (VL) and associated risk factors among Wildlife Soldiers. The study population comprised all 500 Soldiers for Wildlife in Dinder National Park, ensuring comprehensive data collection.

Dinder National Park, located in southeastern Sudan along the border with Ethiopia, serves as a significant study area due to its extensive biodiversity and varied ecosystems. Established in 1935 and covering approximately 10,292 square kilometers, the park is named after the Dinder River, a tributary of the Blue Nile that flows through it. The park features diverse habitats, including woodlands dominated by acacia and other indigenous tree species, extensive grasslands that support a wide range of herbivores, and seasonal wetlands crucial for aquatic species. This rich habitat diversity supports an array of wildlife, including African elephants, lions, leopards, cheetahs, buffalo, giraffes, various antelope species, over 200 bird species, and numerous reptiles and amphibians. The flora of Dinder ranges from dense woodlands to open grasslands and marshy areas, further contributing to its biodiversity [18].

The questionnaire used for data collection was both pre-prepared and pre-tested to ensure its validity and reliability. It contained demographic information and risk factors such as mosquito net usage and awareness regarding control measures of visceral leishmaniasis, as well as self-reported questions based on clinical signs and symptoms of infection.

Data collection involved three primary phases. First, a validated questionnaire was administered to gather detailed information on demographic characteristics and various aspects related to VL. Second, existing medical records were reviewed to identify the prevalence of VL among the soldiers, providing insights into the incidence and management of the disease. Third, an interview was conducted with the Director of the Department of Primary Health Care in Dinder locality to collect data on the VL rate among the soldiers and to assess the quality of health services provided to them. Data were collected from wildlife rangers through face-to-face interviews in a convenient environment to ensure confidentiality.

Data were entered and analyzed using the Statistical Package for Social Science (SPSS), version 24.0. Descriptive statistics summarized the demographic data and prevalence rates, while the associations between different variables were assessed using the Chi-square test, with a p-value of ≤ 0.05 considered statistically significant. Logistic regression was also performed, with calculation of Odds Ratios (OR) and 95% Confidence Intervals (CI). Bivariate analysis (Chi-square test) was conducted at a 5% significance level and 95% confidence interval to examine relationships between the prevalence of VL and risk factors variable, and binary regression (logistic regression) was used to calculate the direction of association between infection and risk factors using OR at 95% CI.

The Odds Ratio and corresponding 95% confidence intervals used in bivariate analysis (as crude odds ratio) and embedded in logistic regression analysis (as adjusted odds ratio) determined relationships between the prevalence of visceral leishmaniasis and several risk factors.

## Ethics Statement

Ethical approval for this research was obtained in three stages. First, approval was secured from the Ministry of Health and Social Development, Sennar State - General Administration of Basic Health Care - Diseases Department – Kala-azar Department in 2/5/2021. Second, it was approved by the Health Services Administration of Al-Dindar Locality, under the number A K S / M D / 50/ 1, 4/5/2021. Finally, the Police Presidency of the Dinder Federal Park, affiliated with the General Administration for Wildlife Protection, headed by the Police Forces - Ministry of Interior, granted approval under the number A A H H B/M D/M1/, 4/5/2021. Additionally, verbal consent was obtained from the Wildlife Soldiers prior to conducting the interviews.

## Result

### Demograhpic characteristics

The study included a total of 500 participants, predominantly male (93%) with only 7% female. The age distribution showed that 13.4% of the participants were under 20 years old, 68% were between 20 and 30 years old, and 18.6% were over 30 years old.

Regarding religious affiliation, 95.4% of the participants were Muslim, while 4.6% identified as non-Muslim. The marital status of the participants was nearly evenly split, with 45.8% married, 45.2% single, 8% divorced, and 1% widowed. In terms of educational level, 12% were illiterate, 14.2% had completed primary school, 60.8% had a high school education, and 13% were graduates. Monthly income levels varied, with 8% of the participants earning a low income, 69.4% earning a medium income, and 22.6% earning a high income. Family size was categorized into three groups: 25.4% had less than five members, 53.6% had between five and eight members, and 21% had more than nine members. Participants’ years of experience in their roles showed that 8.6% had less than one year of experience, 45.8% had between two and five years of experience, and 45.6% had over five years of experience. Regarding military rank, 15.6% of the participants were officers, while the majority, 84.4%, were soldiers. All these demographic characteristics are indicated in the **(Table 1)**

**Table 1:** Demographic characteristics of wildlife soldiers in Dinder National Park, Sudan, (n **= 500).**

| Characteristic | Frequency | % |
| --- | --- | --- |
| **Sex** |  |  |
| Male | 465 | 93 |
| Female | 35 | 7 |
| **Age groups** |  |  |
| <20 | 67 | 13.4 |
| 20 – 30 | 340 | 68 |
| >30 | 93 | 18.6 |
| **Religion** |  |  |
| Muslim | 477 | 95.4 |
| Not Muslim | 23 | 4.6 |
| **Marital status** |  |  |
| Married | 229 | 45.8 |
| Single | 226 | 45.2 |
| Divorce | 40 | 8 |
| Widow | 5 | 1 |
| **Educational level** |  |  |
| Illiterate | 60 | 12 |
| Primary school | 71 | 14.2 |
| High school | 304 | 60.8 |
| Graduate | 65 | 13 |
| **Monthly income** |  |  |
| Low | 40 | 8 |
| Medial | 347 | 69.4 |
| High | 113 | 22.6 |
| **Family size** |  |  |
| <5 | 127 | 25.4 |
| 5 – 8 | 268 | 53.6 |
| >9 | 105 | 21 |
| **Years of Experience** |  |  |
| <1 year | 43 | 8.6 |
| 2 - 5 years | 229 | 45.8 |
| >5 years | 228 | 45.6 |
| **Military rank** |  |  |
| Officer | 78 | 15.6 |
| Soldier | 422 | 84.4 |

### Prevalence of VL and associated factors

The prevalence of visceral leishmaniasis (VL) among the participants was 27.6%, with 138 individuals reported positive, while 72.4%, or 362 individuals, reported negative. **(Figure 1)** Regarding previous VL infections, 7.4% of the participants reported having an infection within the past month, 11% reported an infection within the past year, and 10.6% reported an infection more than a year ago. The majority, 71%, reported no previous VL infection. **(Figure 2)** The study revealed the following associations between various demographic and risk factors with the prevalence of visceral leishmaniasis (VL) **(Table 2)**.

**Figure I:**
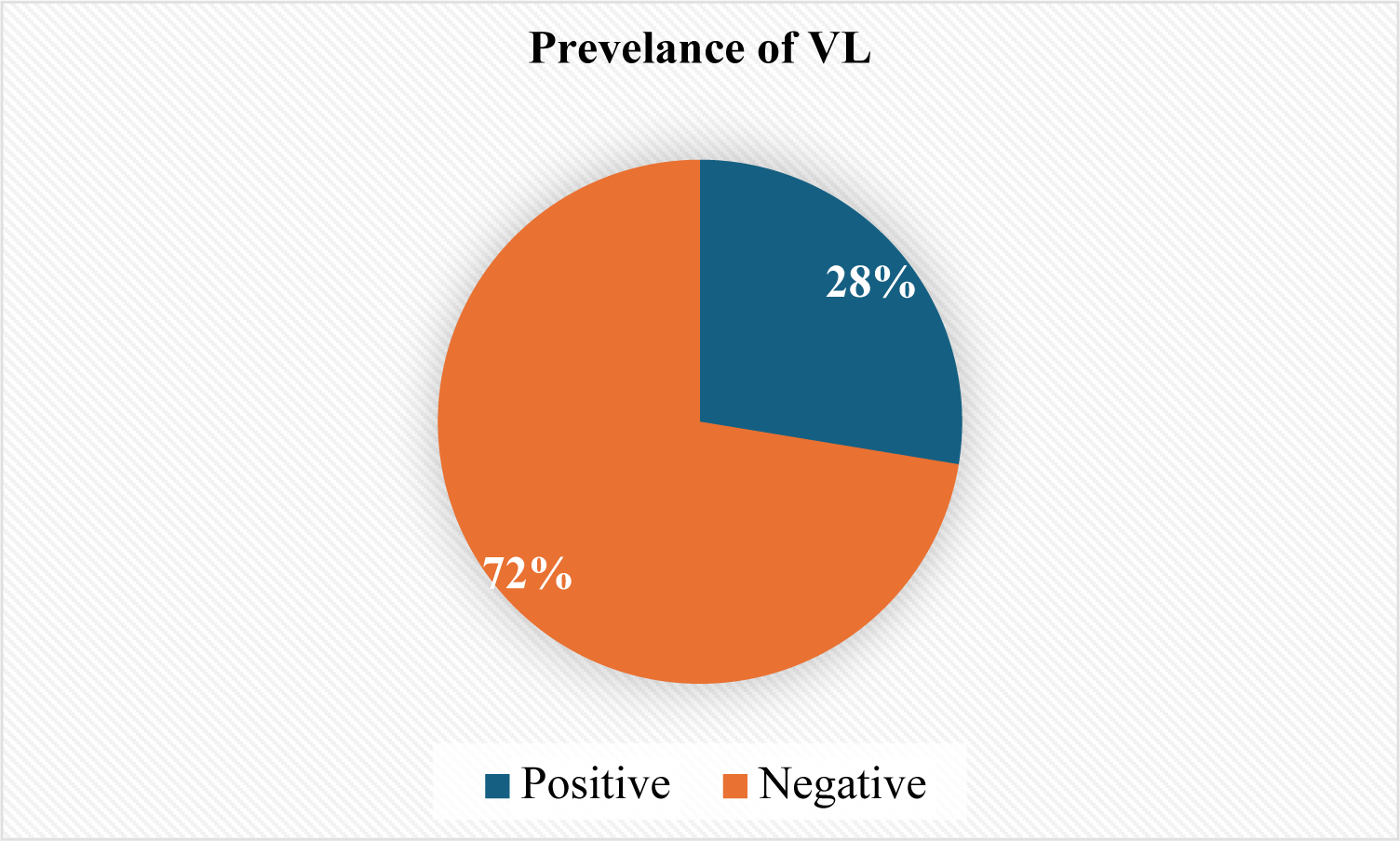
Self-reported Prevalence of VL among wildlife rangers in Dinder National Park, Sudan, (n = 500).

**Figure II:**
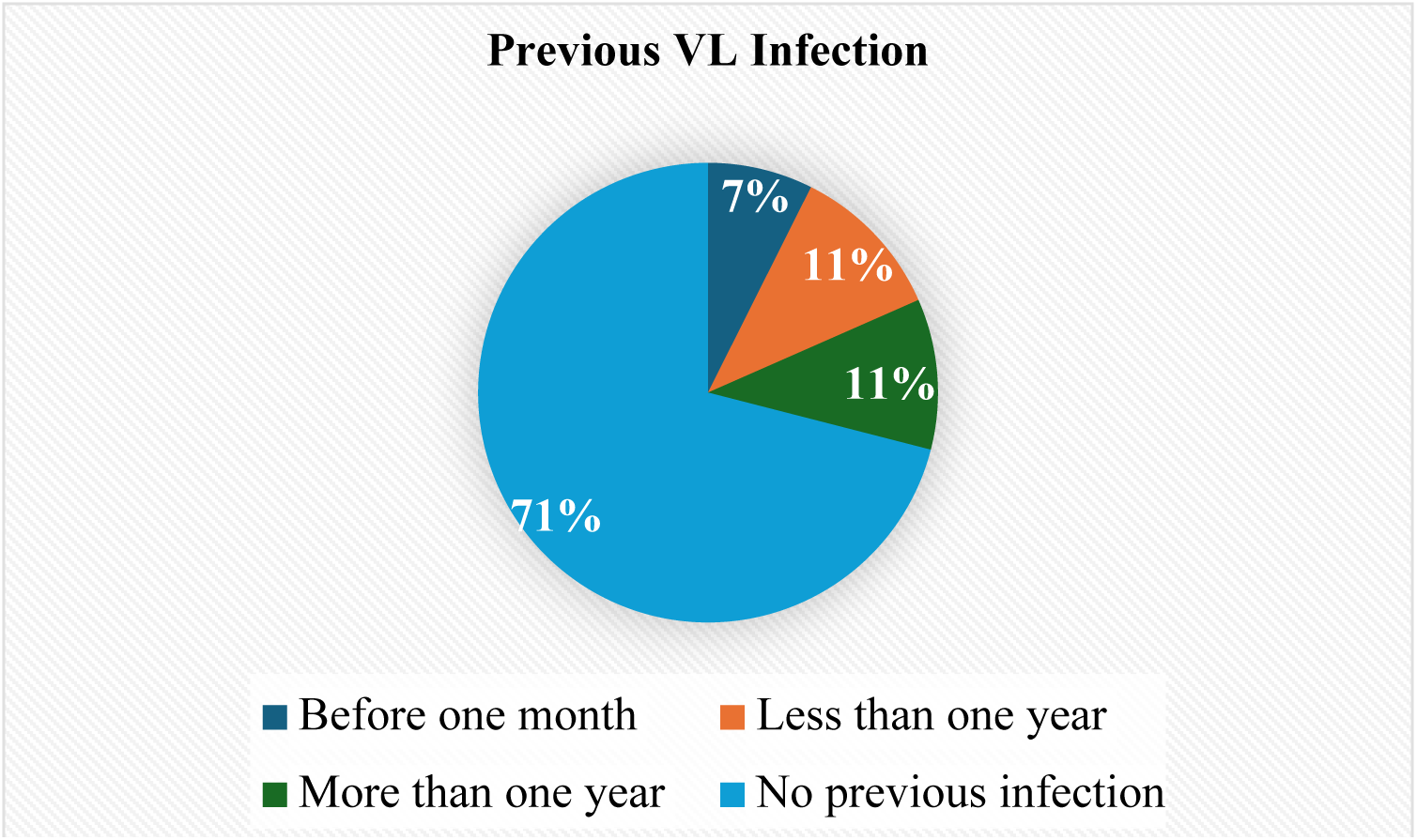
Previous infection with VL among wildlife soldiers in Dinder National Park, Sudan, (n = 500).

**Table 2:** Relationship between prevalence of VL and risk factors among wildlife soldiers in Dinder National Park, Sudan, (n = 500).

| Variable | Category | Positive (%) | Negative (%) | Total (%) | P-value | OR (95% CI) |
| --- | --- | --- | --- | --- | --- | --- |
| Educational Level | Illiterate | 16 (3.20) | 44 (8.80) | 60 (12.00) | 0.001 | -- |
|  | Primary school | 29 (5.80) | 42 (8.40) | 71 (14.20) |  | 1.895 (0.901 - 3.983) |
|  | High school | 67 (13.40) | 237 (47.40) | 304 (60.80) |  | 0.777 (0.413 - 1.462) |
|  | Graduate | 26 (5.20) | 39 (7.80) | 65 (13.00) |  | 1.833 (0.861 - 3.906) |
| Family Income | Low | 14 (2.80) | 26 (5.20) | 40 (8.00) | 0.352 | -- |
|  | Medium | 96 (19.20) | 251 (50.20) | 347 (69.40) |  | 0.71 (0.36 - 1.42) |
|  | High | 28 (5.60) | 85 (17.00) | 113 (22.60) |  | 0.61 (0.28 - 1.33) |
| Military Rank | Officer | 33 (6.60) | 45 (9.00) | 78 (15.60) | 0.004 | -- |
|  | Soldier | 105 (21.00) | 317 (63.40) | 422 (84.40) |  | 0.45 (0.27 - 0.75) |
| Awareness about Signs and Symptoms of VL | Yes | 128 (25.60) | 260 (52.00) | 388 (77.60) | 0.000 | -- |
|  | No | 10 (2.00) | 102 (20.40) | 112 (22.40) |  | 0.20 (0.10 - 0.39) |
| Awareness about VL Transmission Methods | Yes | 120 (24.00) | 255 (51.00) | 375 (75.00) | 0.000 | -- |
|  | No | 18 (3.60) | 107 (21.40) | 125 (25.00) |  | 0.36 (0.21 - 0.61) |
| Awareness about Control of VL | Yes | 124 (24.80) | 307 (61.40) | 431 (86.20) | 0.012 | -- |
|  | No | 14 (2.80) | 55 (11.00) | 69 (13.80) |  | 0.63 (0.34 - 1.18) |
| Sleeping under the Net | Yes | 44 (8.80) | 172 (34.40) | 216 (43.20) | 0.001 | -- |
|  | No | 94 (18.80) | 190 (38.00) | 284 (56.80) |  | 1.93 (1.28 - 2.92) |

Among illiterate participants, 16 individuals (3.20%) reported positive for VL, and 44 (8.80%) reported negative. Participants with a primary school education had 29 individuals (5.80%) test positive and 42 (8.40%) test negative, with an Odds Ratio (OR) of 1.895 (95% Confidence Interval [CI]: 0.901 - 3.983), compared to the illiterate group. Those with a high school education had 67 individuals (13.40%) test positive and 237 (47.40%) test negative, with an OR of 0.777 (95% CI: 0.413 - 1.462). Among graduates, 26 individuals (5.20%) tested positive and 39 (7.80%) tested negative, with an OR of 1.833 (95% CI: 0.861 - 3.906). The p-value for educational level was 0.001, indicating a statistically significant difference.

Participants with a low income had 14 individuals (2.80%) test positive and 26 (5.20%) test negative. Those with a medium income had 96 individuals (19.20%) test positive and 251 (50.20%) test negative, with an OR of 0.71 (95% CI: 0.36 - 1.42). Participants with a high income had 28 individuals (5.60%) test positive and 85 (17.00%) test negative, with an OR of 0.61 (95% CI: 0.28 - 1.33). The p-value for family income was 0.352, indicating no statistically significant difference.

Among officers, 33 individuals (6.60%) tested positive for VL, and 45 (9.00%) tested negative. Soldiers had 105 individuals (21.00%) test positive and 317 (63.40%) test negative, with an OR of 0.45 (95% CI: 0.27 - 0.75). The p-value for military rank was 0.004, indicating a statistically significant difference.

Participants who were aware of the signs and symptoms of VL had 128 individuals (25.60%) test positive and 260 (52.00%) test negative. Those who were not aware had 10 individuals (2.00%) test positive and 102 (20.40%) test negative, with an OR of 0.20 (95% CI: 0.10 - 0.39). The p-value for awareness about signs and symptoms was 0.000, indicating a statistically significant difference.

Participants aware of VL transmission methods had 120 individuals (24.00%) test positive and 255 (51.00%) test negative. Those unaware had 18 individuals (3.60%) test positive and 107 (21.40%) test negative, with an OR of 0.36 (95% CI: 0.21 - 0.61). The p-value for awareness about VL transmission methods was 0.000, indicating a statistically significant difference.

Participants aware of VL control methods had 124 individuals (24.80%) test positive and 307 (61.40%) test negative. Those unaware had 14 individuals (2.80%) test positive and 55 (11.00%) test negative, with an OR of 0.63 (95% CI: 0.34 - 1.18). The p-value for awareness about control of VL was 0.012, indicating a statistically significant difference.

Participants who slept under a net had 44 individuals (8.80%) test positive and 172 (34.40%) test negative. Those who did not sleep under a net had 94 individuals (18.80%) test positive and 190 (38.00%) test negative, with an OR of 1.93 (95% CI: 1.28 - 2.92). The p-value for sleeping under the net was 0.001, indicating a statistically significant difference.

## Interview Results

Interviews were conducted with the Director of the Department of Primary Health Care in Dinder Locality to explore perspectives on the factors contributing to the high prevalence of visceral leishmaniasis among Wildlife Soldiers in Dander, Sinnar State. The 2021 interview results revealed several key insights. According to the Director, Wildlife Soldiers stationed in forested areas are at heightened risk due to the presence of leishmania vectors, increasing their susceptibility to infection compared to other populations. Additionally, many soldiers lack adequate protective equipment such as mosquito nets, and even those who possess them often do not use them consistently. These factors likely contribute to the elevated prevalence of visceral leishmaniasis in the area.

## Discussion

This study investigated the prevalence of visceral leishmaniasis (VL) and associated risk factors among Wildlife Soldiers in Dinder National Park, Sudan. The findings revealed significant associations between VL prevalence and various demographic and risk factors, including educational level, military rank, awareness of VL signs and symptoms, awareness of VL transmission methods, awareness of VL control measures, and the practice of sleeping under a mosquito net. The prevalence of VL in this study was 27.6%, which is alarmingly high compared to global averages, particularly in regions outside of endemic areas. This high prevalence aligns with findings from studies conducted in other parts of Sudan and East Africa, where environmental conditions and socio-economic factors contribute to higher rates of infection. A recent systematic review by [10] in Sudan revealed that the overall pooled prevalence of human leishmaniasis in the country is 21%. Cutaneous leishmaniasis (CL) is the most common type, with a prevalence of 26%, followed by visceral leishmaniasis (VL) at 18%. Notably, the prevalence of VL reported in [10] review is lower than that observed in our current study. The review also indicates that central Sudan has the highest prevalence of human leishmaniasis at 27%, which aligns with the findings of our current study in the same region. However, the overall prevalence of human leishmaniasis appears to be decreasing over time [10]. Another study by [16], conducted in Eastern Gedaref State, Sudan, among children, reported that 47 out of 145 suspected cases (32%) were identified as having VL. This prevalence is higher than that among adults, indicating that children are more likely to contract VL. The study also found a significant association between rural residence, male gender, and VL among children. This suggests that rural areas are higher-risk zones due to socioeconomic conditions, poor housing, and inadequate sanitary conditions. Similarly, in the same eastern region, [17] reported that VL represents a significant health burden in the villages of Eastern Gedaref State, being one of the major causes of death in the area. The study conducted by [19] investigated the clinical and demographic features of visceral leishmaniasis (VL) in an African population. The findings indicate that males are at a higher risk than females, with the majority of patients being children under 15 years old. A similar study [20] conducted in Gadaref, Eastern Sudan, also reported that children and men are at higher risk of visceral leishmaniasis (VL). This increased risk among children is attributed to their tendency to play outside, making them more likely to be exposed to the sandfly vectors that transmit the Leishmania parasites. For men, the higher risk is due to their likelihood of working in outdoor environments, such as farming and forestry, where they are more frequently exposed to sandflies. The association between educational level and VL prevalence was significant, with illiterate participants having a higher prevalence of infection (3.20%). This finding suggests that education plays a crucial role in disease prevention and awareness. Similar results have been reported in other studies, where higher educational levels correlated with lower prevalence rates of various infectious diseases, including VL. The ORs indicate that primary school and graduate participants had increased odds of VL compared to illiterate participants, though not statistically significant. Family income was not significantly associated with VL prevalence in this study. However, those with low and medium incomes had higher prevalence rates compared to those with high income. This finding is consistent with previous research indicating that lower socio-economic status is often linked to higher rates of VL due to limited access to healthcare and preventive measures. The lack of statistical significance might be due to the relatively small sample size or other confounding factors not accounted for in this study. Military rank showed a significant association with VL prevalence, with soldiers having a higher prevalence (21.00%) compared to officers (6.60%). This disparity may be attributed to differences in living conditions, exposure to infected sandflies, and access to protective measures. Similar patterns have been observed in other occupational health studies, where lower-ranking personnel often face greater exposure to occupational hazards. Awareness about the signs and symptoms of VL, transmission methods, and control measures was significantly associated with lower prevalence rates. Participants who were aware of these aspects had a much lower prevalence of VL. This finding underscores the importance of health education and awareness campaigns in controlling the spread of VL. Previous studies have also highlighted the critical role of awareness and education in reducing the incidence of VL and other vector-borne diseases. The practice of sleeping under a mosquito net was significantly associated with lower VL prevalence. Participants who did not sleep under a net had nearly twice the odds of contracting VL compared to those who did. This finding aligns with extensive research demonstrating the effectiveness of insecticide-treated nets in reducing the transmission of vector-borne diseases, including malaria and VL.

## Conclusion

This study highlights the critical need for targeted public health interventions to reduce the prevalence of VL among Wildlife Soldiers in Dinder National Park. Key strategies should include educational programs to raise awareness about VL, its transmission, and preventive measures, along with efforts to improve living conditions and access to protective measures such as mosquito nets. Addressing these factors could significantly reduce the burden of VL in this high-risk population.

## Implications for Future Research

The findings of this study highlight several key areas for future research to further understand and mitigate the burden of visceral leishmaniasis (VL) among Wildlife Soldiers in Dinder National Park, Sudan.

Detailed epidemiological studies are needed to track the incidence and prevalence of VL over time, identifying seasonal variations and long-term trends. These studies should explore the role of environmental factors specific to Dinder National Park, such as climate patterns, wildlife interactions, and habitat characteristics, in influencing VL transmission dynamics.

Research into socio-economic and behavioral factors is crucial. This includes investigating the socio-economic determinants of health that contribute to VL risk, focusing on the impacts of poverty, education, and access to healthcare services. Assessing the effectiveness of current health education programs and developing targeted interventions to improve awareness and preventive behaviors among different demographic groups within the soldier population is also important.

Intervention strategies should be evaluated to determine the effectiveness of various preventive measures, such as insecticide-treated nets, personal protective equipment, and environmental management, in reducing VL transmission. Community-based intervention trials should be implemented and assessed to determine the best practices for VL control in the context of Wildlife Soldiers and other at-risk populations in similar settings.

The accessibility and quality of healthcare services available to Wildlife Soldiers need to be examined, focusing on diagnostic capabilities, treatment availability, and overall healthcare infrastructure. Qualitative research is necessary to understand the barriers to healthcare access and utilization, including cultural beliefs, stigma, and logistical challenges.

Molecular and genetic studies can provide valuable insights into the specific strains of Leishmania parasites circulating in the region and their potential resistance to standard treatments. Exploring the genetic susceptibility of the local population to VL, identifying potential genetic markers that could inform personalized treatment and prevention strategies, is also important.

Occupational health and safety research should investigate the specific conditions that increase exposure to sandflies and other vectors associated with wildlife conservation work. Developing and evaluating occupational health and safety programs tailored to the unique needs and risks faced by Wildlife Soldiers is crucial.

Collaboration between local, national, and international health organizations should be fostered to share knowledge, resources, and best practices for VL control. Policy changes that support the allocation of resources to VL research, prevention, and treatment programs should be advocated for ensuring sustained efforts to combat the disease.

By addressing these research areas, future studies can provide a more comprehensive understanding of VL in Dinder National Park and contribute to the development of effective, evidence-based interventions to reduce the burden of this disease among Wildlife Soldiers and other vulnerable populations.

## Limitations

This study, while providing valuable insights into the prevalence of visceral leishmaniasis (VL) and associated risk factors among Wildlife Soldiers in Dinder National Park, Sudan, has several limitations that should be considered when interpreting the results.

First, the cross-sectional design of the study limits the ability to establish causal relationships between identified risk factors and VL prevalence. While associations can be observed, it is not possible to determine the direction of these relationships or infer causality.

Second, the reliance on self-reported data for certain variables, such as awareness of VL signs and symptoms and use of mosquito nets, may introduce reporting bias. Participants might over-report positive behaviors or under-report negative behaviors due to social desirability bias.

Third, the study’s focus on Wildlife Soldiers may limit the generalizability of the findings to other populations. The unique environmental and occupational conditions faced by Wildlife Soldiers in Dinder National Park might not reflect those experienced by the general population or other occupational groups in Sudan or other regions.

Fourth, the study did not account for potential confounding variables that could influence the observed associations. Factors such as nutritional status, other underlying health conditions, and genetic predispositions were not controlled for, which may affect the study’s findings.

Fifth, the sample size, while comprehensive for the Wildlife Soldiers in Dinder National Park, may still be limited for certain subgroup analyses. Small sample sizes within specific categories, such as those with low educational levels or low income, might reduce the statistical power to detect significant differences.

Sixth, the use of a single diagnostic method for VL prevalence may not capture all cases accurately. Misclassification of cases, either false positives or false negatives, could affect the prevalence estimates and the strength of associations with risk factors.

Seventh, the study did not include detailed environmental and ecological data, which are crucial for understanding the transmission dynamics of VL. Factors such as vector density, habitat characteristics, and climatic conditions were not assessed, limiting the ability to fully understand the environmental determinants of VL risk.

Lastly, the study period and timing might influence the results. Seasonal variations in sandfly activity and VL transmission could affect prevalence rates, and the study might not capture these temporal dynamics accurately.

Acknowledging these limitations is essential for contextualizing the findings and guiding future research efforts to address these gaps and build a more comprehensive understanding of VL in this high-risk population.

## Data Availability

The data that support the findings of this study are available from the corresponding author upon reasonable request.

## Funding

This research received no specific grant from any funding agency in the public, commercial, or not-for-profit sectors.

## Competing interests

The authors declare that they have no competing interests.

## Authorship contributions

**BI** and **ME** contributed to the conception of the work. **ME, HS, MGAA**, and **BI** were involved in drafting the article, data analysis, and critical revision of the article. **AE** gave final approval of the version to be published. All authors read and finalized the article and agreed to its publication.

## Notes

### Competing Interest Statement

The authors have declared no competing interest.

### Funding Statement

The author(s) received no specific funding for this work.

